# A cross-sectional study of the association between frequency of telecommuting and unhealthy dietary habits among Japanese workers during the COVID-19 pandemic

**DOI:** 10.1101/2021.06.28.21259558

**Authors:** Yoshiko Kubo, Tomohiro Ishimaru, Ayako Hino, Masako Nagata, Kazunori Ikegami, Seiichiro Tateishi, Mayumi Tsuji, Shinya Matsuda, Yoshihisa Fujino, for the CORoNaWork project

**Affiliations:** Department of Environmental Epidemiology, Institute of Industrial Ecological Sciences, University of Occupational and Environmental Health, Japan; Department of Mental Health, Institute of Industrial Ecological Sciences, University of Occupational and Environmental Health, Japan; Department of Occupational Health Practice and Management, Institute of Industrial Ecological Sciences, University of Occupational and Environmental Health, Japan; Department of Work Systems and Health, Institute of Industrial Ecological Sciences, University of Occupational and Environmental Health, Japan; Department of Occupational Medicine, School of Medicine, University of Occupational and Environmental Health, Japan; Department of Environmental Health, School of Medicine, University of Occupational and Environmental Health, Japan; Department of Preventive Medicine and Community Health, School of Medicine, University of Occupational and Environmental Health, Japan

**Author notes:** **Correspondence to** Yoshihisa Fujino, M.D., M.P.H., Ph.D., Department of Environmental Epidemiology, Institute of Industrial Ecological Sciences, University of Occupational and Environmental Health, Japan, 1-1, Iseigaoka, Yahatanishiku, Kitakyushu, 807-8555, Japan. **Author contributions:** YK: writing of the manuscript; TI: creating the questionnaire, review of the manuscript, and advice on interpretation; AH, MN, and KI: review of the manuscript and advice on interpretation; ST, MT, and SM: review of the manuscript, advice on interpretation, and funding for the research; YF: overall survey planning, creating the questionnaire, analysis, and drafting of the manuscript.

**Keywords:** Teleworking, COVID-19, Feeding behavior, Breakfast, Meal Times, Japan

## Abstract

2.

**Objective:** Due to the COVID-19 pandemic, telecommuting has become a new way of working that has not only changed individuals’ work, but also their health and lifestyle. We examined the relationship between telecommuting frequency and unhealthy dietary habits among Japanese workers.

**Methods:** A total of 33,302 workers completed an Internet survey about telecommuting and dietary habits. Data from 13,468 office workers who telecommuted were analyzed. Telecommuting frequency during the COVID-19 pandemic was extracted from a questionnaire. The odds ratios (ORs) of four types of dietary habits, namely, skipping breakfast, solitary eating, lower meal frequency, and meal substitution associated with telecommuting frequency were estimated using multilevel logistic regression nested in the prefecture of residence to control for differences in residential area.

**Results:** The multivariate OR of skipping breakfast was 1.15 (95% CI: 1.03–1.29, p=0.013) for participants who telecommuted in excess of four days per week compared to those who rarely telecommuted. Similarly, the OR of solitary eating, lower meal frequency and meal substitution were 1.44 (95%CI: 1.28–1.63, p<0.001), 2.39 (95%CI: 1.66–3.44, p<0.001), and 1.26 (95%CI: 1.04–1.51, p=0.015) for those who telecommuted in excess of four days per week compared to those who rarely telecommuted. There was a statistically significant increase in the dose-response trend in ORs of solitary eating (p for trend <0.001), lower meal frequency (p for trend <0.001), and meal substitution (p for trend =0.001) with increasing telecommuting frequency.

**Conclusion:** Telecommuters may develop unhealthy dietary habits, indicating the need for strategies to help telecommuters manage their nutrition and diet.

## Introduction

Telecommuting is a symbol of the new normal brought about by the global COVID-19 pandemic. Governments around the world have recommended telecommuting as a key measure to restrict the movement of people in an effort to contain the spread of infection.^1^ In Japan, many companies have introduced telecommuting at the request of government directives.^2^ Although some companies had implemented telecommuting before the COVID-19 pandemic, current telecommuting practices differ from those initiated before the pandemic in many respects: the practice advanced rapidly, including among companies and workers who had no experience with telecommuting; was not based on workers’ preference; has been taken up by workers in many types of jobs; and has been introduced even in small and medium-sized companies. As a result, the impact of telecommuting on workers’ lives and society has been enormous.

For many workers, telecommuting is a new way of working that has not only changed their work, but also their health, lifestyle, family and community relationships. However, the full impact of telecommuting is not well understood. The change to telecommuting has had an impact on health problems previously identified in office workers, such as back pain, visual display terminal-related symptoms, and eye strain, as well as work productivity.^3,4^ There are also reports that telecommuting affects stress, communication, and mental health.^5^ In addition, telecommuting is thought to be having a significant effect on lifestyle habits like alcohol consumption, exercise, and sleep.^6–10^

Telecommuting is also expected to significantly change workers’ eating habits.^9,11–13^ For example, reduced commuting leaves more time for other life activities and preparations; lower frequency of going out and less physical activity due to telecommuting may lead to lower calorie consumption; and more flexible use of time at home may lead to more snacking. Government directives to refrain from going out to prevent the spread of COVID-19 may also have reduced the frequency of eating out and shopping for food. Such changes brought about by telecommuting will thus likely affect how frequently individuals eat meals, whether or not they skip breakfast, the types of foods they consume as meals, and who they eat with. A systematic review reported that there is an association between dietary habits and various occupational factors, including work hours, shift work and job stress.^14^ However, few studies have examined how workers’ dietary habits have changed due to telecommuting in the COVID-19 pandemic, when they have been required to refrain from going out.^11^

The present study examined four dietary changes, namely skipping breakfast, solitary eating, meal substitution, and lower meal frequency, based on the general recommendations of Japan’s National Public Health Policy, Health Japan 21. According to the policy, skipping breakfast can lead to unbalanced nutrient intake.^15,16^ Further, solitary eating, lack of variety in food intake, and consumption of instant foods can lead to potential health problems. One goal of Health Japan 21 is to promote “eating at least one proper meal a day with at least two family members for at least 30 minutes.” Previous studies indicated that skipping breakfast and lower meal frequency are linked to health conditions such as overweight, obesity, cardiovascular disease, type 2 diabetes, and high cholesterol.^17–20^

We hypothesized that skipping breakfast, solitary eating, meal substitution, and lower meal frequency are associated with telecommuting. We thus examined the relationship between the frequency of telecommuting and unhealthy dietary habits among Japanese workers during the COVID-19 pandemic.

## Methods

### Study Design and Participants

A cross-sectional Internet monitor study was conducted between December 22 and 26, 2020, while Japan was experiencing its third wave of COVID-19 infection. Details of the study protocol are published elsewhere.^21^ Briefly, the target participants were those who were employed and aged between 20 and 65 years at the baseline survey. Given that COVID-19 infection rates in Japan vary according to region, we used regional sampling to select participants based on region, job type, and sex. The regions were categorized into five groups depending on the regional infection level. Participants were selected to balance the sample among the five regions, sexes, and occupations. The survey was conducted by Cross Marketing Inc. (Tokyo, Japan), which has 4.7 million registered monitors. A total of 605,381 monitors were sent an email to invite them to participate in a study. Of these, 55,045 answered the initial screening questions, and 33,302 who satisfied criteria for worker status, region, sex, and age completed to the survey.

Of the 33,302 participants, we excluded 6,266 who provided fraudulent responses, which were defined as follows: extremely short response time (≤6 minutes), extremely low body weight (<30 kg), extremely short height (<140 cm), inconsistent answers to similar questions (e.g., inconsistent answers to questions about marital status and living area), and incorrect answers to a question purposely included to detect fraudulent responses (choose the third largest number from the following five numbers). A total of 13,468 (6,896 men and 6,572 women) of the remaining 27,036 participants who stated that they predominantly perform desk work were included in the analysis.

This study was approved by the ethics committee of the University of Occupational and Environmental Health, Japan (reference No. R2-079 and R3-006). Participants provided informed consent by completing a form on the survey website.

### Frequency of telecommuting

Telecommuting frequency was determined using the question, “How frequently do you currently telecommute?” Participants chose from the following options: “more than 4 days per week,” “more than 2 days per week,” “less than 1 day per week,” and “hardly ever.”

### Dietary habits

The following questions were used to determine whether participants missed breakfast and ate alone: “How often do you usually eat breakfast?” and “How often do you eat all of the meals in a day alone?” Respondents chose from the following five options: “6–7 days per week,” “4–5 days per week,” “2–3 days per week,” “less than 1 day per week,” and “almost never.”

We also inquired about the number of meals and the number of alternative meals eaten per day to determine meal frequency and adoption of meal substitution using the following questions: “How often do you consume meals a day?” and “How many times a day do you eat only individual food items and packaged foods (yogurt, protein bars, pastries, etc.)?” Respondents chose from the following five options: “more than 4 times per day,” “3 times a day,” “2 times a day,” “1 time a day,” and “hardly ever.”

### Other covariates

We controlled for the following confounding variables: age, sex, education, equivalent income (household income divided by the square root of household size), cohabitation status, smoking status (never; quit smoking more than one year ago; quit smoking within the past year; started smoking less than one year ago; smoking for more than one year), alcohol consumption (6–7 days a week; 4–5 days a week; 2–3 days a week; less than 1 day a week; hardly ever), and physical activity more than 30 minutes per day (6–7 days a week; 4–5 days a week; 2–3 days a week; less than 1 day a week; hardly ever).

Additionally, the community-level variable was the cumulative COVID-19 infection rate in the participants’ prefecture of residence one month before the start of the survey.

### Statistical analysis

We determined the age-sex-adjusted and multivariate-adjusted odds ratios (ORs) of unfavorable dietary habits associated with telecommuting frequency. We defined unfavorable dietary habits as skipping breakfast, solitary eating, lower meal frequency, and meal substitution. To control for differences in residential area, we used a multilevel logistic model nested in the prefecture of residence. The multivariate model included sex, age, equivalent household income, education, cohabitation status, smoking status, alcohol consumption, and physical activity. Additionally, we used the COVID-19 infection rate in each prefecture as a prefecture-level variable in all comparisons. To avoid multicollinearity between marital status and cohabitation status, only cohabitation status was included in the multivariate analysis. We also examined the interaction between telecommuting and age, sex, and living alone on dietary habit. A p value <0.05 was considered statistically significant. Further, we performed Bonferroni correction to control for false positives. All analyses were conducted using Stata (Stata Statistical Software: Release 16; StataCorp LLC, TX, USA).

## Results

Participants’ characteristics by telecommuting frequency are summarized in Table 1. Of the 13,468 office workers included in the analysis, approximately 30% telecommuted at least once a week. Those who telecommuted more often tended to be slightly older and male, and less likely to be living with family. Those who telecommuted more frequently tended to have more unfavorable eating habits. Among workers who hardly telecommuted, 25.5% missed breakfast, 25.9% ate all meals alone, 1% ate less than two meals a day, and 6.6% adopted meal substitution. The corresponding proportions among workers who telecommuted in excess of four days per week were 28.7%, 37.0%, 2.5%, and 8.4%, respectively.

**Table 1.** Subjects’ characteristics according to frequency of telecommuting

| Factor | Frequency of telecommuting |  |  |  |
| --- | --- | --- | --- | --- |
|  | ≥4 days/week<br>n=2042 | 2-3 days/week<br>n=1058 | ≤1 day/week<br>n=952 | Hardly ever<br>n=9416 |
| Age, mean (SD) | 49.1 (10.0) | 48.4 (10.3) | 48.1 (10.2) | 47.3 (10.2) |
| Sex, male (%) | 1159 (56.8%) | 591 (55.9%) | 613 (64.4%) | 4533 (48.1%) |
| Marriage status, married | 991 (48.5%) | 650 (61.4%) | 625 (65.7%) | 5498 (58.4%) |
| Cohabitation | 1512 (74.0%) | 828 (78.3%) | 770 (80.9%) | 7532 (80.0%) |
| Equivalent income (million JPY) |  |  |  |  |
| 40-249 | 490 (24.0%) | 115 (10.9%) | 95 (10.0%) | 1652 (17.5%) |
| 250-375 | 433 (21.2%) | 214 (20.2%) | 176 (18.5%) | 2593 (27.5%) |
| 376-499 | 440 (21.5%) | 274 (25.9%) | 241 (25.3%) | 2456 (26.1%) |
| ≥500 | 679 (33.3%) | 455 (43.0%) | 440 (46.2%) | 2715 (28.8%) |
| Education |  |  |  |  |
| Junior high school | 20 (1.0%) | 8 (0.8%) | 3 (0.3%) | 58 (0.6%) |
| High school | 346 (16.9%) | 111 (10.5%) | 129 (13.6%) | 2343 (24.9%) |
| University, graduate school,<br>vocational school, junior college | 1676 (82.1%) | 939 (88.8%) | 820 (86.1%) | 7015 (74.5%) |
| Current smoker | 500 (24.5%) | 267 (25.2%) | 261 (27.4%) | 2206 (23.4%) |
| Alcohol consumption |  |  |  |  |
| 6-7 days a week | 458 (22.4%) | 251 (23.7%) | 230 (24.2%) | 1967 (20.9%) |
| 4-5 days a week | 167 (8.2%) | 123 (11.6%) | 94 (9.9%) | 691 (7.3%) |
| 2-3 days a week | 252 (12.3%) | 139 (13.1%) | 148 (15.5%) | 1177 (12.5%) |
| Less than 1 day a week | 353 (17.3%) | 192 (18.1%) | 190 (20.0%) | 1603 (17.0%) |
| Hardly ever | 812 (39.8%) | 353 (33.4%) | 290 (30.5%) | 3978 (42.2%) |
| Physical activity more than 30 minutes per day |  |  |  |  |
| 6-7 days a week | 158 (7.7%) | 58 (5.5%) | 73 (7.7%) | 402 (4.3%) |
| 4-5 days a week | 138 (6.8%) | 86 (8.1%) | 60 (6.3%) | 474 (5.0%) |
| 2-3 days a week | 263 (12.9%) | 165 (15.6%) | 158 (16.6%) | 1033 (11.0%) |
| Less than 1 day a week | 306 (15.0%) | 186 (17.6%) | 161 (16.9%) | 1270 (13.5%) |
| Hardly ever | 1177 (57.6%) | 563 (53.2%) | 500 (52.5%) | 6237 (66.2%) |
| Miss breakfast less than 3 days per week | 586 (28.7%) | 264 (25.0%) | 230 (24.2%) | 2405 (25.5%) |
| Eat all meals alone at least 4 days a week | 755 (37.0%) | 327 (30.9%) | 262 (27.5%) | 2437 (25.9%) |
| Consume less than two meals per day | 52 (2.5%) | 17 (1.6%) | 13 (1.4%) | 90 (1.0%) |
| Eat only individual food items or packaged food more than twice a day | 171 (8.4%) | 102 (9.6%) | 70 (7.4%) | 618 (6.6%) |

The ORs of the four unhealthy dietary habits associated with telecommuting frequency are summarized in Table 2. The multivariate OR of skipping breakfast was 1.15 (95% CI: 1.03–1.29, p=0.013) among participants who telecommuted in excess of four days per week compared to those who rarely telecommuted. Similarly, the multivariate ORs of solitary eating, lower meal frequency, and meal substitution were 1.44 (95%CI: 1.28–1.63, p<0.001), 2.39 (95%CI: 1.66–3.44, p<0.001), and 1.26 (95%CI: 1.04–1.51, p=0.015), respectively. There was a statistically significant increasing dose-response trend in ORs of solitary eating (Bonferroni-adjusted p for trend <0.001), lower meal frequency (Bonferroni-adjusted p for trend <0.001), and meal substitution (Bonferroni-adjusted p for trend =0.004) with increasing telecommuting frequency.

**Table 2.** Odds ratios of unhealthy dietary habit associated with frequency of telecommuting

| Outcome variable |  | Frequency of telecommuting |  |  |  |  |  |  |  |  |  | p for trend<sup>c</sup> |
| --- | --- | --- | --- | --- | --- | --- | --- | --- | --- | --- | --- | --- |
|  |  | Hardly ever<br>n=9416 | ≤1 day/week<br>n=952 |  |  | 2-3 days/week<br>n=1058 |  |  | ≥4 days/week<br>n=2042 |  |  |  |
|  |  |  | OR | 95%CI | p | OR | 95%CI | p | OR | 95%CI | p |  |
| Miss breakfast more than 3 days per week |  |  |  |  |  |  |  |  |  |  |  |  |
|  | model 1<sup>a</sup> | reference | 0.89 | 0.76-1.04 | 0.142 | 0.90 | 0.77-1.05 | 0.183 | 1.13 | 1.02-1.27 | 0.026 | 0.992 |
|  | model 2<sup>b</sup> | reference | 0.95 | 0.81-1.12 | 0.529 | 0.97 | 0.83-1.13 | 0.686 | 1.15 | 1.03-1.29 | 0.013 | 0.272 |
| Eat all meals alone at least 4 days a week |  |  |  |  |  |  |  |  |  |  |  |  |
|  | model 1<sup>a</sup> | reference | 1.03 | 0.88-1.20 | 0.743 | 1.09 | 0.95-1.26 | 0.220 | 1.52 | 1.37-1.68 | <0.001 | <0.001 |
|  | model 2<sup>b</sup> | reference | 1.07 | 0.90-1.27 | 0.459 | 1.16 | 0.98-1.36 | 0.081 | 1.44 | 1.28-1.63 | <0.001 | <0.001 |
| Consume less than two meals per day |  |  |  |  |  |  |  |  |  |  |  |  |
|  | model 1<sup>a</sup> | reference | 1.48 | 0.82-2.68 | 0.189 | 1.75 | 1.03-2.97 | 0.040 | 2.89 | 2.03-4.12 | <0.001 | <0.001 |
|  | model 2<sup>b</sup> | reference | 1.35 | 0.74-2.45 | 0.332 | 1.52 | 0.88-2.63 | 0.130 | 2.39 | 1.66-3.44 | <0.001 | <0.001 |
| Eat only individual food items or packaged food more than twice a day |  |  |  |  |  |  |  |  |  |  |  |  |
|  | model 1<sup>a</sup> | reference | 1.14 | 0.88-1.47 | 0.330 | 1.56 | 1.24-1.95 | <0.001 | 1.37 | 1.14-1.64 | 0.001 | <0.001 |
|  | model 2<sup>b</sup> | reference | 1.10 | 0.85-1.44 | 0.466 | 1.46 | 1.16-1.84 | 0.001 | 1.26 | 1.04-1.51 | 0.015 | 0.004 |

<sup>a</sup> Model 1 was adjusted for age and sex.<sup>b</sup> Model 2 was adjusted for age, sex, education, annual equivalent household income, cohabitation, smoking status, alcohol consumption, and physical activity.<sup>c</sup> Bonferroni-adjusted p for trend

Multivariate adjustment slightly attenuated the results of the age-adjusted model, but did not affect the findings. Multivariate adjustment also revealed a marginal dose-response trend (p for trend=0.068) in the association between skipping breakfast and telecommuting frequency that was not observed in the univariate analysis (p=0.248).

In addition, we found that there was no significant effect of the interaction between telecommuting and age or sex on dietary habit. In contrast, the interaction between living alone and telecommuting had a significant effect (p<0.001) on solitary eating. Table 3 shows the ORs of solitary eating associated with telecommuting frequency stratified by living arrangement. While there was no association between telecommuting and solitary eating among workers who lived with their families, telecommuting frequency among workers who lived alone was associated with eating alone. Compared to those who did not telecommute, the OR of solitary eating was 1.4 (95%CI: 1.01–1.93, p=0.042) among those who telecommuted two or more days per week, and 2.18 (95%CI: 1.68–2.82, p<0.01) among those who telecommuted four or more days per week.

**Table 3.**
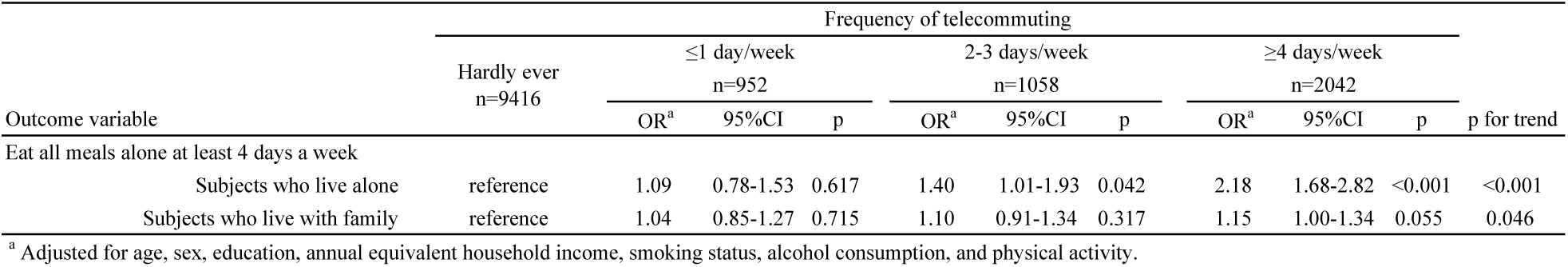
Odds ratios of solitary eating associated with frequency of telecommuting stratified by living arrangement

## Discussion

We found that telecommuting frequency was associated with four unhealthy dietary habits: skipping breakfast, solitary eating, lower meal frequency, and meal substitution. To our knowledge, this is the first study to examine these associations in the COVID-19 pandemic, during which telecommuting has been recommended around the world to prevent the spread of infection.

Eating breakfast is widely recommended for maintaining good health.^22,23^ Numerous studies have demonstrated an association between skipping breakfast and adverse health effects such as overweight, obesity, cardiovascular disease, type 2 diabetes, and poor cognitive performance.^17–19^ In this study, participants who worked from home in excess of four or more a week were significantly more likely to skip breakfast compared to those who rarely worked from home. However, contrary to our hypothesis, there was no clear dose-response relationship between telecommuting frequency and frequency of skipping breakfast. Several reasons may explain the lack of a dose-response relationship. First, families need breakfast regardless of a worker’s telecommuting status. This is supported by the fact that adjustment for cohabitation status in the multivariate analysis produced a dose-response trend that approached significance. Second, the definition of breakfast is ambiguous.^25^ While some people define breakfast as the first meal eaten upon waking, others define it as any meal eaten in the morning. Thus, vagueness and misclassification of definitions may have affected the observed associations.

We also found that greater telecommuting frequency was associated with more frequent solitary eating. In Japanese culture, avoidance of solitary eating is considered an important public health practice.^26^ One of the goals of Japan’s national public health policy is to promote community eating.^15^ The declining birthrate, women’s participation in society, and fragmentation of family life in Japan are thought to have led to an increase in solitary eating, which is linked to unbalanced nutrition, disrupted life rhythm, and loneliness.^28^ However, there is little formal evidence on the health effects of solitary eating, with very few studies having examined the impact of solitary eating on health.^29,30^ During the COVID-19 pandemic, in addition to urging the public to stay home, the government also suggested that individuals avoid eating with people with whom they do not normally spend time. Frequent telecommuting likely further reduced individuals’ opportunity to share meals with others. Given the surging public health challenges associated with loneliness and mental health around the world brought about by the pandemic,^20^ our results suggest the need to care for those who are forced to eat alone while telecommuting.

There is currently no consensus on how many meals a person should consume per day for good health. However, studies have shown that, in general, consuming more frequent meals per day is associated with a lower risk of weight gain, obesity, high cholesterol, and type 2 diabetes.^19,20^ The number of meals consumed per day differs by country and time. Japanese people generally eat three times a day. The current standard health guidance in Japan recommends balanced nutrition and necessary calorie intake at breakfast, lunch, and dinner, and refraining from placing extreme restrictions on the number of meals consumed per day.^31^ The present study showed that an increase in telecommuting frequency was associated with an increase in the number of people who ate fewer meals per day. We speculate that there may be several possible explanations for this finding. Increased telecommuting frequency may decrease the need for people to go out for commuting purposes and thus their physical activity. This would reduce calorie consumption, which may lead to lower appetite and less frequent meals. Workers who telecommute may be more likely to spend longer hours at home and snack, which may compensate for the food that would normally be consumed at meals. Requests to refrain from going out to prevent the spread of infection may have led people to refrain from going out for meals and shopping, thus reducing the number of meals eaten. Alternatively, telecommuting may be affecting individuals’ loneliness and mental health, thus affecting their dietary patterns.^31–33^

Additionally, we found that increased telecommuting frequency was associated with more frequent meal substitution with individual food items and processed or packaged foods such as bread, cookies, nutritional supplements, yogurt, and pastries. Eating a few individual items and packaged foods may compensate for the consumption of fewer meals. Directives to refrain from going out are likely to have led individuals to eating meals they can easily prepare and buy food with a longer shelf life. The health effects of such meal substitution are unknown, as they depend on the food and frequency of consumption.

This study showed that telecommuters were more likely to have an unhealthy diet, which is associated with the risk of cardiovascular disease and diabetes.^34^ Telecommuters are also reportedly less physically active.^35,36^ Together, these results indicate the importance of managing cardiovascular and diabetes risk in telecommuters. While guidelines for the work environment have been published for desk workers who telecommute, including specifications for room illumination, display illumination, room temperature, and ergonomic desks and chairs, ^34^ strategies for promoting healthy diet and exercise habits among telecommuters form a new challenge in occupational health practice.

There are several limitations to this study. First, while we found an association between poor dietary habits and telecommuting frequency, these relationships may not be solely related to the impact of telecommuting, but the COVID-19 pandemic at large and a variety of other consequences such as staying at home and reduced interaction with people. Second, because this was a cross-sectional study, we cannot prove causality. However, we believe that it is logically unlikely that dietary habits could directly affect telecommuting. Third, we examined both the frequency of telecommuting and eating habits in the questionnaire. Dietary surveys are associated with several well-known disadvantages, including under-reporting, over-reporting, and bias.^35,36^ In addition, we did not specify a time period in our questions. Because of the inter-day variability in dietary intake, the impact of these factors on our results is unclear.^37^ Fourth, the effects of potentially important confounding factors that were not examined in this study are unclear. In particular, bedtime and waking time, which are thought to be affected by telecommuting, may be associated with skipping breakfast. However, if the change in bedtime and waking time due to telecommuting causes individuals to skip breakfast, then bedtime and waking time are pathways rather than confounders; adjusting for these factors would lead to over-adjustment. Additionally, we found that adjusting for changes in sleep duration (increase, no change, or decrease) due to telecommuting had no effect on the results.

In conclusion, we found that increasing telecommuting frequency was associated with an increasing number of unhealthy dietary habits. This study suggests that in addition to challenges related to physical activity, loneliness, and mental health, telecommuters may also be developing unhealthy dietary habits, indicating the need for strategies to help telecommuters manage their nutrition and diet.

**Figure 1.**
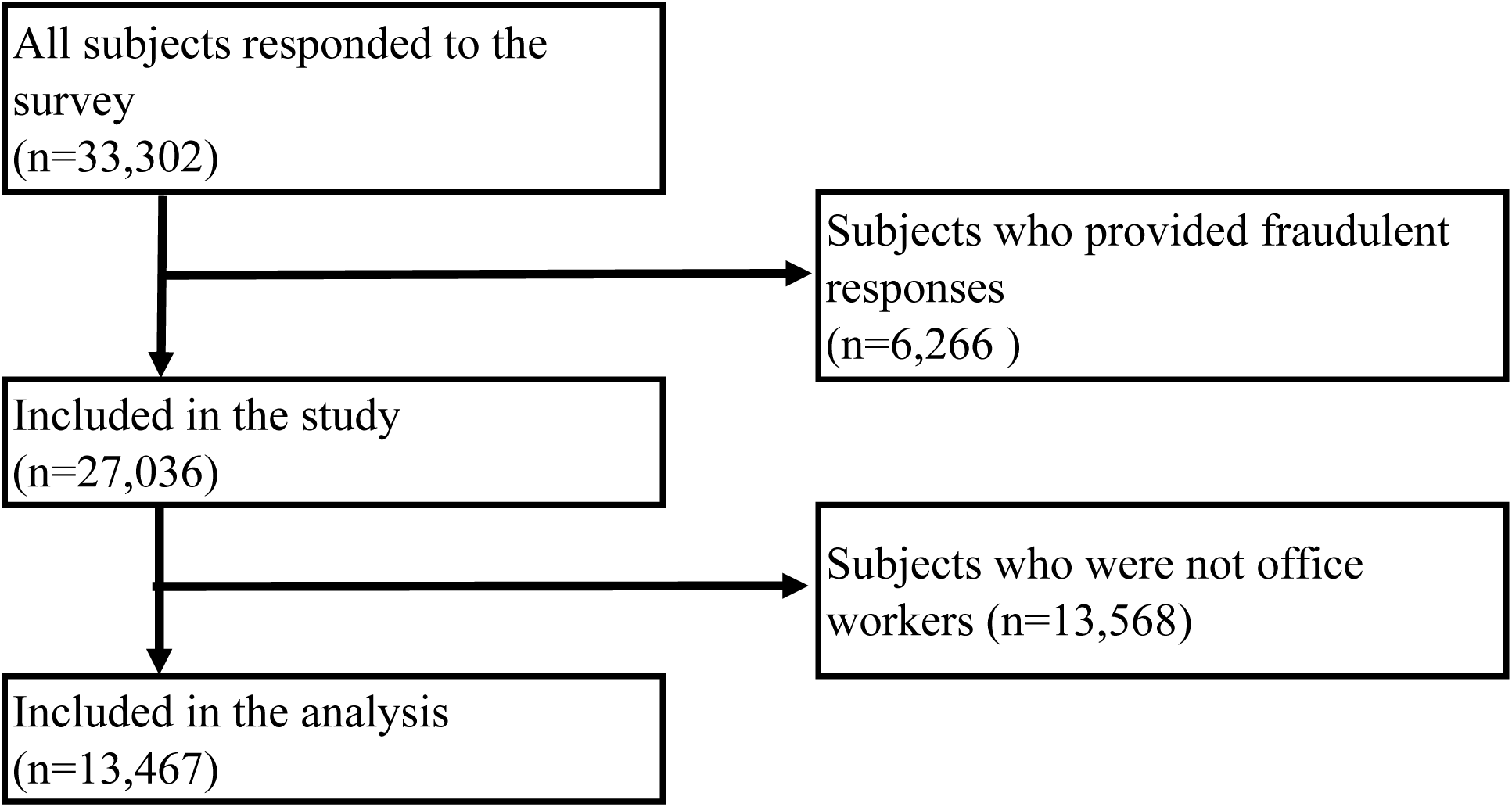
Flow chart of study population.

## Data Availability

The datasets generated and/or analyzed during the current study are not publicly available due to ethical restrictions, but are available from the corresponding author on reasonable request.

## 4. Acknowledgements

This study was supported and partly funded by the University of Occupational and Environmental Health, Japan; General Incorporated Foundation (Anshin Zaidan); The Development of Educational Materials on Mental Health Measures for Managers at Small-sized Enterprises; Health, Labour and Welfare Sciences Research Grants; Comprehensive Research for Women’s Healthcare (H30-josei-ippan-002); Research for the Establishment of an Occupational Health System in Times of Disaster (H30-roudou-ippan-007); scholarship donations from Chugai Pharmaceutical Co., Ltd., the Collabo-Health Study Group, and Hitachi Systems, Ltd.

The current members of the CORoNaWork Project, in alphabetical order, are as follows: Dr. Yoshihisa Fujino (present chairperson of the study group), Dr. Akira Ogami, Dr. Arisa Harada, Dr. Ayako Hino, Dr. Hajime Ando, Dr. Hisashi Eguchi, Dr. Kazunori Ikegami, Dr. Kei Tokutsu, Dr. Keiji Muramatsu, Dr. Koji Mori, Dr. Kosuke Mafune, Dr. Kyoko Kitagawa, Dr. Masako Nagata, Dr. Mayumi Tsuji, Ms. Ning Liu, Dr. Rie Tanaka, Dr. Ryutaro Matsugaki, Dr. Seiichiro Tateishi, Dr. Shinya Matsuda, Dr. Tomohiro Ishimaru, and Dr. Tomohisa Nagata. All members are affiliated with the University of Occupational and Environmental Health, Japan.

## 5. Disclosures

### Ethical approval

This study was approved by the Ethics Committee of the University of Occupational and Environmental Health, Japan (reference No. R2-079 and R3-006).

### Informed consent

Informed consent was obtained via a form on the survey website.

### Registry and the Registration No. of the study/Trial

N.A.

### Animal Studies

N.A.

